# Utility of a new lower limb and trunk functional evaluation for pitchers with focus on the physical characteristics of players with a throwing disorder

**DOI:** 10.1101/2023.03.30.23287922

**Authors:** Tomoyuki Matsui, Yoshikazu Azuma, Machiko Hiramoto, Kazuya Seo, Tetsuya Miyazaki, Kanta Matsuzawa, Noriyuki Kida, Toru Morihara

**Affiliations:** Marutmachi Rehabilitation Clinic, Kyoto, Japan; Rehabilitation Unit, University Hospital, Kyoto Prefectural University of Medicine, Kyoto, Japan; Department of Biotechnology, Graduate School of Science and Technology, Kyoto Institute of Technology, Kyoto, Japan

**Keywords:** Throwing disorder, evaluation, pitching, range of motion, rotation

## Abstract

**Objectives:** A pitching motion involves three-dimensional whole body movement. Proper pelvic and trunk rotation movement are important for the prevention of throwing injuries. Given that throwing is not a simple rotation movement, the evaluation of proper motion should reflect muscle strength as well as coordination and pitching motion characteristics. We have devised a throwing rotational assessment (TRA) as a new evaluation of the total rotation angle required for throwing. The purpose of this study was to examine the characteristics of players with throwing disorders compared to a pain-free group using TRA.

**Materials and methods:** The subjects consisted of 164 high school baseball pitchers who participated in a medical check. Pain-induced tests included an elbow hyperextension test and an intra-articular shoulder impingement test. Pitchers who felt pain in either test were classified into a disorder group (n=61). With the subjects in a position similar to the foot contact phase of throwing, the rotation angles of the pelvis and trunk were measured. All tests were performed in the throwing and opposite directions.

**Results:** The disorder group had significantly lower average rotation angles of the pelvis and trunk in the throwing direction and the rotation angle of the trunk in the opposite direction compared to the healthy group.

**Conclusion:** TRA reflects the complex whole body rotation movement. Reductions in rotational angles as assessed in TRA may be associated with throwing disorders. TRA is a simple method that may be useful in the early detection of a throwing disorder and could be used in the systematic evaluation during a medical check, as well as during self-check in the sports field.

## Introduction

In the execution of a throwing motion with a high degree of performance, many factors— such as the range of motion, muscle strength, timing of muscle contractions, and lower limb/trunk/upper limb kinetic chains—are involved [1]. Furthermore, three-dimensional movements (sagittal plane, coronal plane, horizontal plane) are required, with the trunk and pelvis rotation playing a particularly important role in movements along the horizontal plane. Studies of the relationship between rotation during the throwing motion and performance have shown that there is a correlation between the rotation angular velocity of the trunk in the throwing direction and speed [2]. In addition, decreased pelvic rotation velocity [3] and poor pelvic and trunk rotation timing [4] cause a decrease in throwing velocity.

With regard to throwing disorders, research has shown that limitations in trunk rotation affect the shoulder and elbow joints [5]. If there is excessive rotation of the trunk (the body rotates to face forward) immediately before foot contact, the external rotation (valgus) torque in the elbow joint increases [6]. Poor timing in the pelvic and trunk rotation increases shoulder internal rotation torque [7]. Moreover, limitations in the range of motion of the lower limbs and trunk, as well as decreased stability and support, cause impingement of the rotator cuff and labrum in the late cocking phase due to an excessive external rotation and horizontal abduction of the shoulder joint of the throwing arm [8, 9]. Additional research has shown that a glenohumeral internal rotation deficit (GIRD) increases the risk of shoulder and elbow joint disorders four-fold [10]. However, some reports indicate that there is no relationship between the range of motion and ulnar collateral ligament damage in the elbow joints [11]. As such, there is currently no consensus on this issue.

Rather than considering the range of motion of each individual joint, it is necessary to consider a composite of factors including muscle strength and flexibility as a whole, in order to make an objective evaluation of the rotational motion of the entire body. Analyses of the throwing motion using a three-dimensional motion analysis system [2, 6] have shown that it is possible to calculate the detailed joint angle and joint torque, and is a useful method in the evaluation of performance and disorder prevention. However, due to the cost and the time required to perform such an analysis, it is not feasible in a clinical setting for most athletes.

In this study, we recreated the foot contact phase of the throwing motion and devised a throwing rotational assessment (TRA) that indicates the rotation motion of the entire body and the composite elements, which can easily be utilized in clinical settings. The objective of this study was to investigate the reliability and usefulness of TRA in elucidating throwing disorders.

## Subjects and methods

### Subjects

Baseball skill camps are held annually for all high school baseball clubs in Kyoto Prefecture during the off-season. Study participants were recruited at these camps between 2010 and 2011 (Table 1)

**Table 1.** The baseline characteristics of participants.

|  | Data presented as mean ± standard deviation |
| --- | --- |
| Age (years) | 1.63 ± 0.6 |
| Height (cm) | 173.2 ± 5.4 |
| Weight (kg) | 64.5 ± 7.0 |
| Years of baseball experience (years) | 7.7 ± 1.9 |

We enrolled 164 male pitchers, aged 16 to 17 years old (mean, 16.3±0.6 years).

Prior to the start of this study, descriptions of the study were provided to all subjects and their parents/guardians and consent was obtained from all subjects. This study was performed in accordance with the declaration of Helsinki after obtaining approval from the ethics committee at our institution (Raku-gaku-Rin-01-000100).

## Methods

### Diagnosis of the disorder group

An orthopedic surgeon examined the subjects’ shoulder and elbow joints. Players who were determined to require secondary observation in the hospital were placed into the disorder group. All others were placed into the healthy group. Those players who experienced limitations in movement due to leg pain or lower back pain were excluded.

### TRA movement method

TRA of the foot contact phase of the throwing motion (Fig 1) was investigated by a group of three physiotherapists.

**Fig 1.**
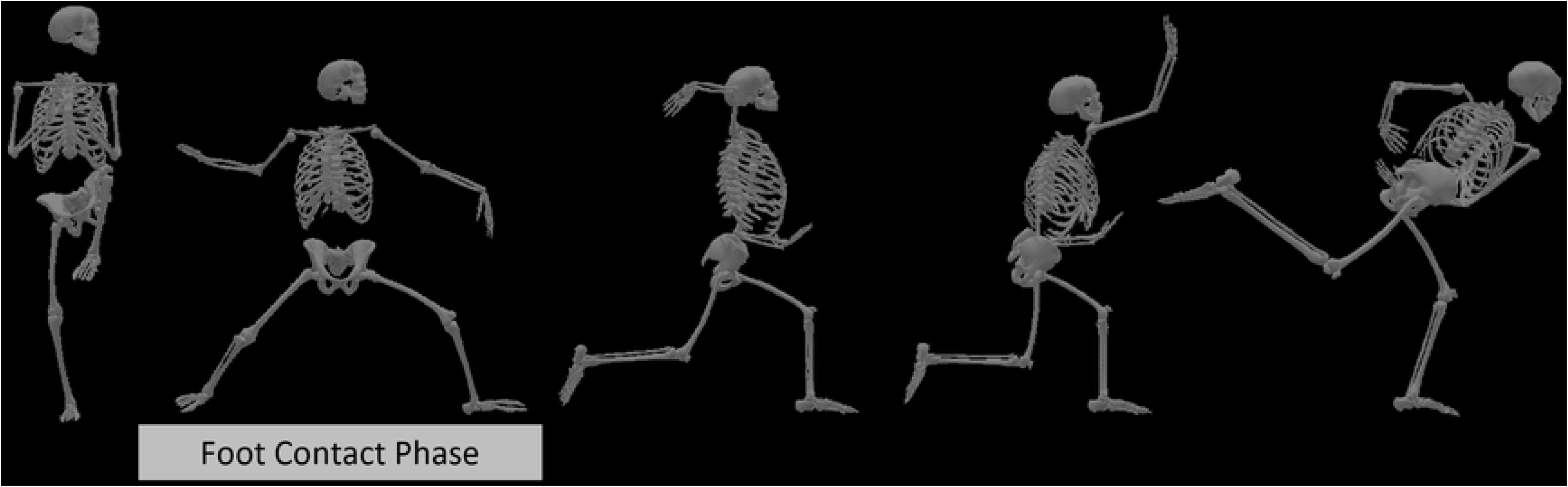
Phase of throwing motion.

The foot contact phase is depicted in relation to other phases of the throwing motion.

The physician performing the examinations and the physiotherapists were blind to the participant’s group status.

The subjects were asked to stand with their feet the same distance apart as when throwing.

Their lower limb on the stance leg side was placed in a position that abducted the hip joint, and the subject took a stance that was neutral between internal and external rotation with their knee joint extended. The lower limb on their lead leg side was positioned so that the hip and knee joints were flexed and the hip joint was abducted and externally rotated. Both lower limbs bore the same amount of weight. From this initial posture, the participant’s hips and thoraxes rotated actively in either the throwing direction or in the opposite direction from the throwing direction.

The angle of the hip rotation was calculated by placing step and horizontal lines from the stationary arm and the movable arm so that it connected both posterior superior iliac spines on either side. We also designated the step direction and horizontal line between the stationary arm and a line connecting the inferior angles of the scapulae on both sides as the movable arm in order to calculate the trunk rotation angle (Fig 2).

**Fig 2.**
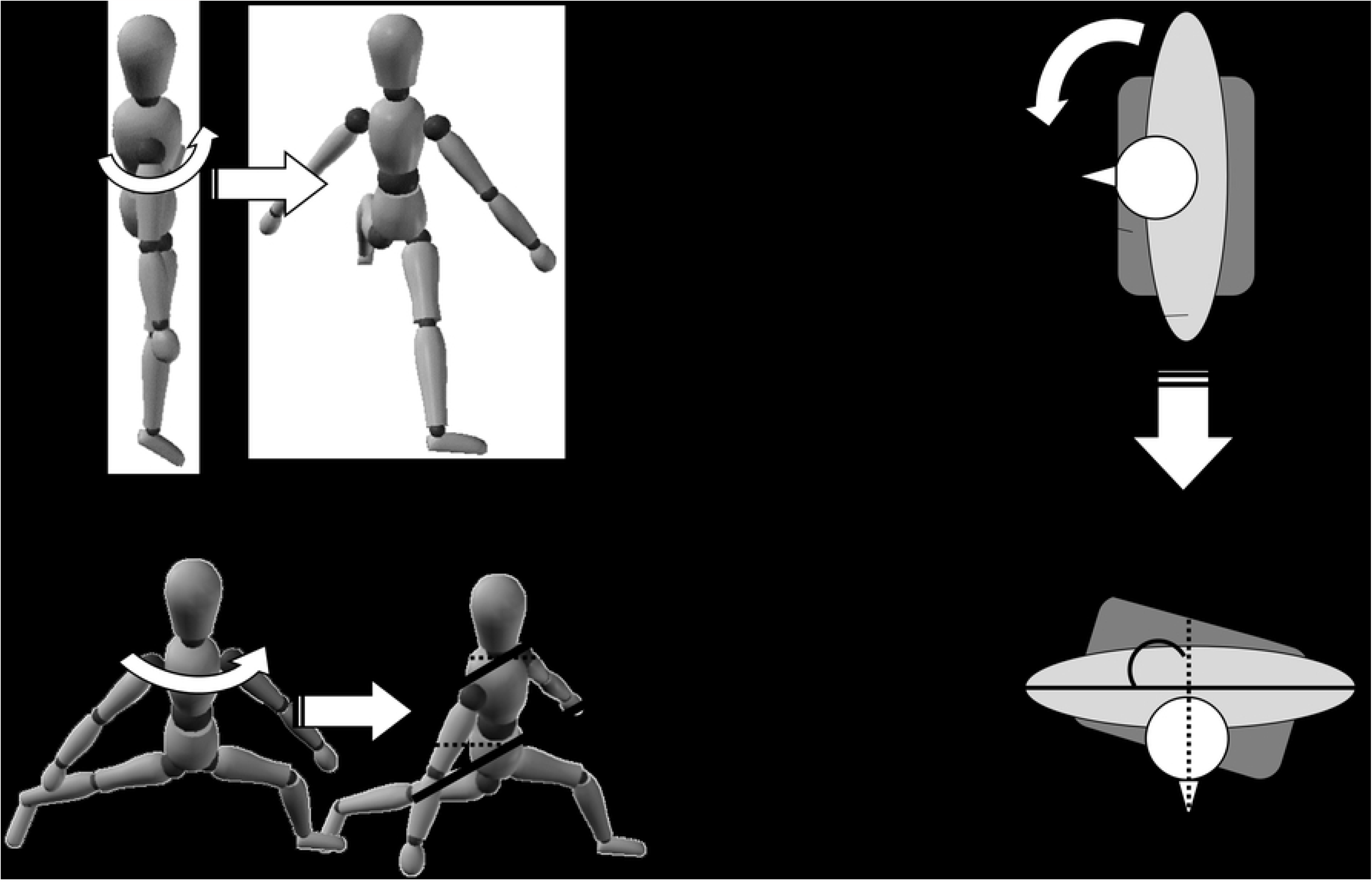
Rotation during the foot contact phase.

In our evaluation of the foot contact phase, the step width was equivalent to actual pitching. The knee joint on the pivoting side is in an extended position and the step side is flexed. The weight bearing is equal for both legs. The pelvis and thorax are rotated in the throwing direction and in the opposite direction.

A standard goniometer was used to measure the angles, which were measured in one-degree increments.

In order to prevent trick motion by the feet, lower legs, or via flexion or lateroflexion of the hip or trunk, a researcher held the lower limb on the stance leg side in place. Visual observations were also made by all the researchers to ensure that compensatory movements did not affect the assessments.

## Statistical analysis

The average pelvic and thoracic angle of rotation was calculated and expressed as the average ± standard deviation. A two-way factorial analysis of variance (ANOVA) with the presence vs. absence of a disorder as the intra-subject factor and throwing vs opposite direction as the intra-subject factor. The disorder group was further subdivided into a shoulder joint disorder group and an elbow joint disorder group and an additional one-way analysis of variance on the resulting three groups (shoulder, elbow, healthy) was performed. Statistical significance was set to p≤0.05.

In order to investigate the utility of TRA as a screening method for throwing disorders, a receiver operating-characteristic (ROC) curve was created for items that showed a significant difference in our comparison of the healthy group and disorder groups. After determining that the area under the curve (AUC) was valid, sensitivity and specificity were calculated as well as the cutoff value using the Youden index (sensitivity + specificity - 1).

In order to investigate the reliability of the measurements, we calculated the intra-class correlation coefficient (ICC) to assess intra-subject reliability (1, 1) and inter-subject reliability (2, 1), as well as the measurement error for the 20 healthy subjects.

## Results

### Detailed description case of the disorders

A total of 26 players were determined by the physician to have shoulder joint disorders and a total of 35 players were determined to have elbow joint disorders (Table 2).

**Table 2.**
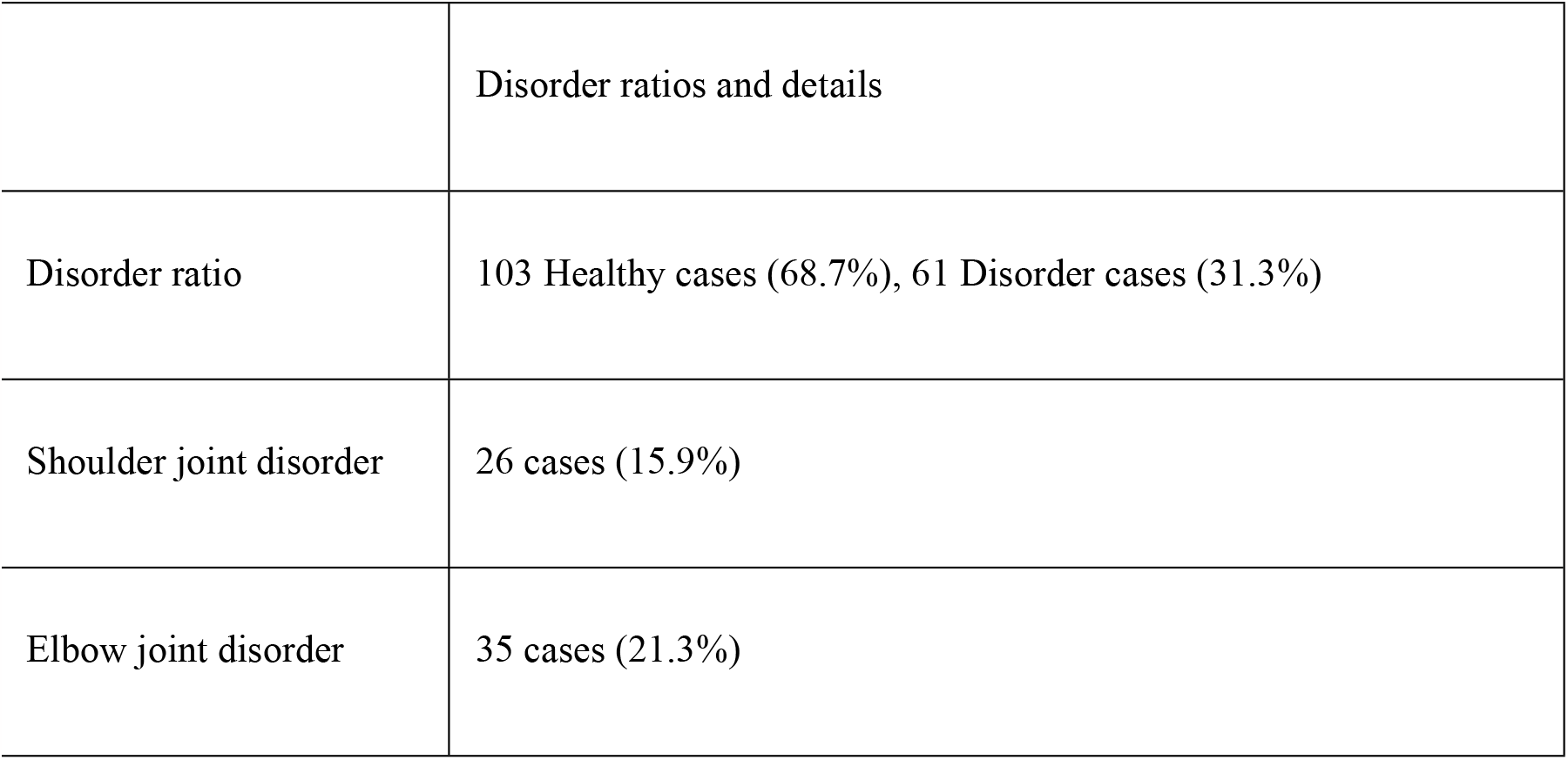
Disorder ratios and details.

All the players underwent follow-up examinations including X-rays, CT scan or MRI at a hospital, whereupon, they were diagnosed with throwing shoulder or throwing elbow disorder as determined by the examination findings.

### Pelvic and thoracic angle of rotation

Since we observed a significant interaction for the pelvic rotation angle in the 2-way ANOVA (Table 3), we tested the simple main effects for movement direction.

**Table 3.**
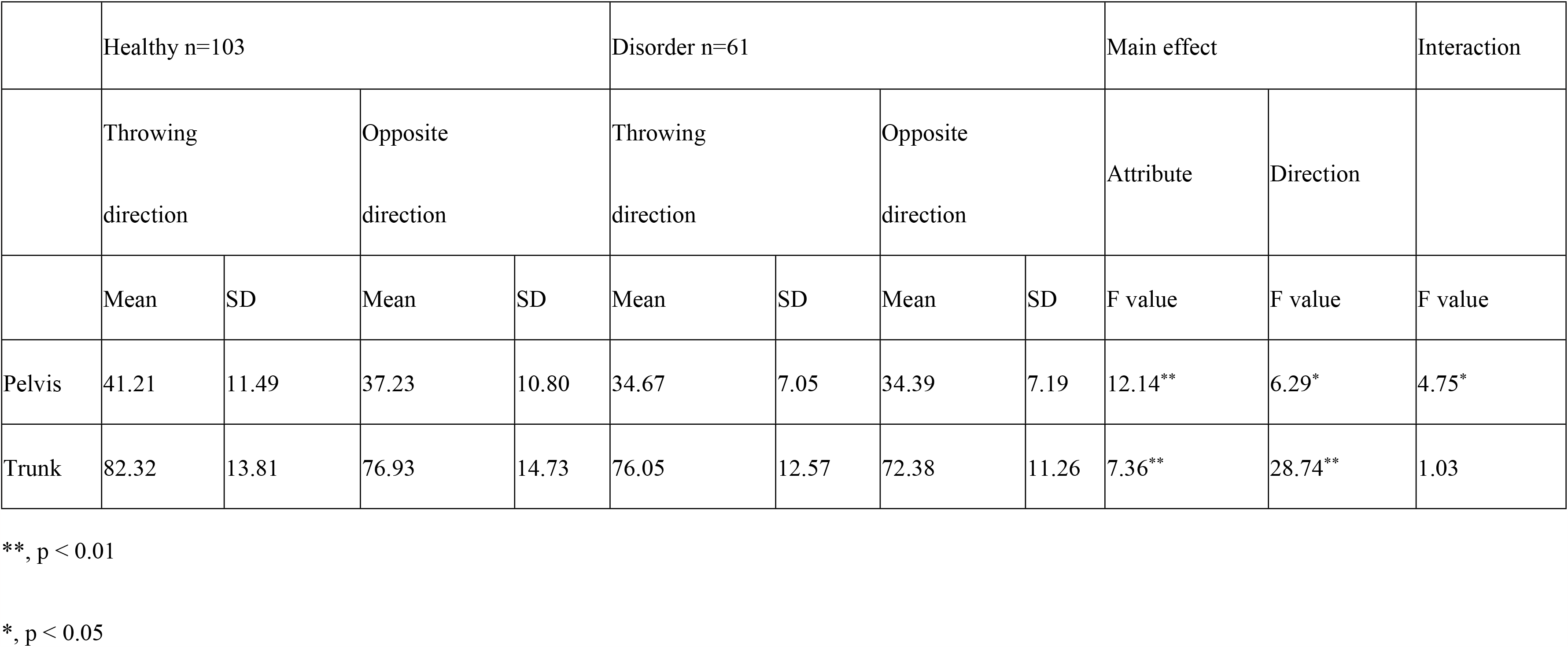
Pelvic and trunk rotation angles.

The results indicated that subjects in the healthy group had significantly higher values for rotation in the throwing direction compared to the opposite direction, but that there were no significant differences in the disorder group. Moreover, the average pelvic rotation angle was significantly higher in the healthy group.

No significant interaction for thoracic rotation angle was observed. However, the healthy group had significantly higher values than those in the disorder group. The comparison of shoulder and elbow joint disorders with the healthy group indicated significantly higher pelvic rotation angle in the healthy group compared to both throwing disorders. In addition, the thoracic rotation angle was significantly higher in the healthy group compared to the elbow joint disorder group (Table 4).

**Table 4.** Pelvic and trunk rotation angles.

|  | Healthy<br>(n=103) |  | Shoulder joint disorder<br>(n=26) |  | Elbow joint disorder<br>(n=35) |  | Main effect |
| --- | --- | --- | --- | --- | --- | --- | --- |
|  | Mean | SD | Mean | SD | Mean | SD |  |
| Pelvis | 41.21 | 11.49 | 33.65 | 7.61 | 35.29 | 6.64 | 8.19 |
| Trunk | 82.32 | 13.81 | 76.54 | 14.86 | 75.69 | 10.77 | 4.22 |

\*\* p < 0.01
\* p < 0.05

### Cutoff values

The cutoff value calculated from the ROC curve was 34.5° for the pelvic angle, with sensitivity of 74.8%, specificity of 52.5%, and an AUC of 0.69 (Fig 3).

**Fig 3.**
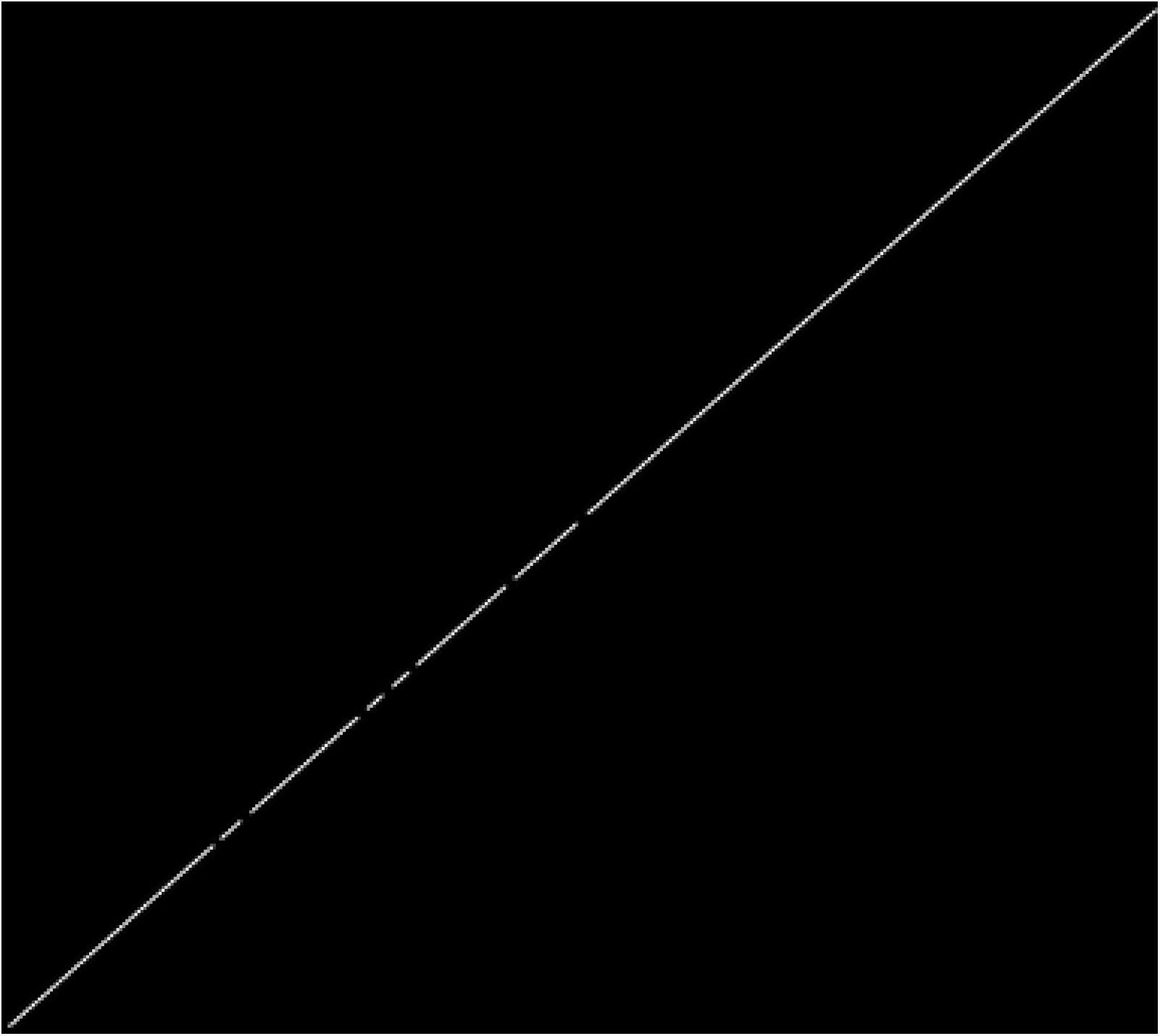
ROC curve with pelvic angle for throwing disorders.

AUC, area under the curve

The cutoff value calculated from the ROC curve for the thoracic angle was 81°, with sensitivity of 66.0%, specificity of 52.4%, and an AUC of 0.67 (Fig 4).

**Fig 4.**
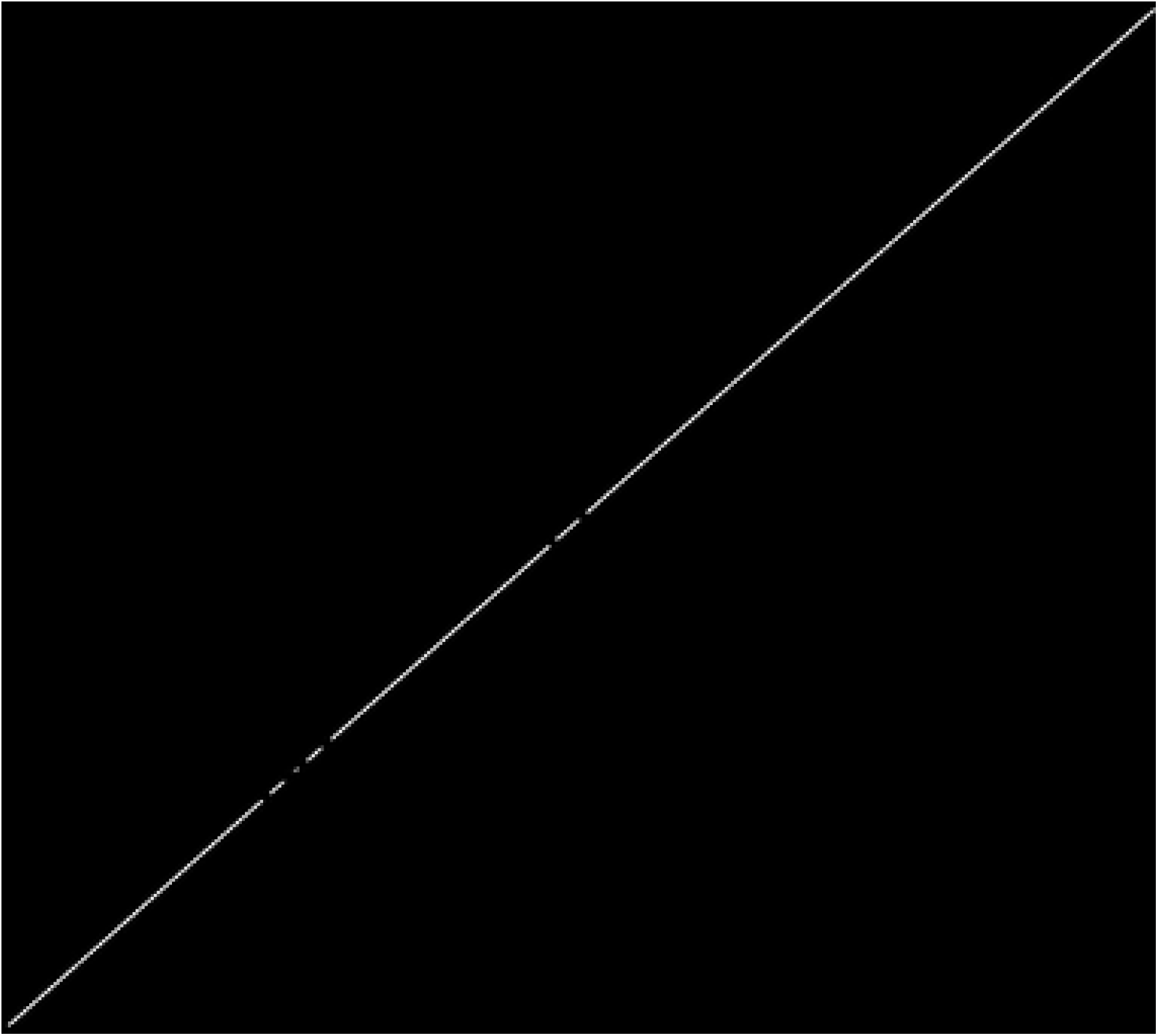
ROC curve with thoracic angle for throwing disorders.

AUC, area under the curve

## Reliability

ICC (1, 1) was 0.92 for the pelvis and 0.94 for the thorax. ICC (2, 1) was 0.82 for the pelvis and 0.85 for the thorax, indicating high reliability. Measurement error was 2.27° for the pelvis and 2.46° for the thorax.

## Discussion

Common throwing disorders involving the shoulders include proximal humerus epiphysiolysis in elementary and junior high school students who have not yet undergone epiphyseal closure, and internal impingement syndrome in high school and university students who have undergone epiphyseal closure [8, 12]. Internal impingement syndrome is a common throwing shoulder disorder and was first reported by Walch in 1992. It is characterized by excessive abduction and horizontal external rotation of the throwing shoulder joint during the late cocking phase of the throwing motion, which causes impingement of the rotator cuff and glenoid rim within the shoulder joint [9]. If the impingement is repeated with continuous throwing, it may lead to lateral rotator cuff tear and damage to the glenoid rim. Thus, early detection and prevention are important.

The prevalence of posterior and lateral elbow joint disorders is high. Posterior disorders are divided into the following three types: posteromedial elbow impingement (malacia, synovitis, and other types of early osteoarthrosis due to sports, olecranon spur), incomplete olecranon epiphyseal closure, and olecranon stress fracture [13-15]. The mechanism of onset it thought to be a valgus extension overload from the late cocking phase to the acceleration phase and a mechanical door stop action in the follow-through phase. Internal disorders are caused by excessive external rotation of the elbow during the acceleration phase and traction stress on the inner tissue. These disorders are highly prevalent, with 70% of pitchers experiencing pain on the inner side [16].

Risk factors for throwing disorder include the number of pitches, the type of pitches, the height of the pitcher, and the pitching form [17, 18]. Pitching form in particular is affected by physical functions such as the range of motion and muscle strength [19]. Investigation into differences for the range of motion for the right and left sides in baseball pitchers has elucidated that the shoulder on the pitching side has an increased external rotation angle and decreased internal rotation angle due to bone-related factors such as reduced flexibility of the posterior tissue of the shoulder, elongation of the anterior capsule of the shoulder, and increased humeral retroversion angle [20-24]. In addition, investigation into lower limb and trunk range of motion has shown that in healthy junior high and high school baseball pitchers, the rotation angle of the neck and trunk was significantly larger and that during hip joint internal rotation, the angle on the lead leg side was significantly larger than on the stance leg side [25]. The TRA used in this study also indicated that there was significant rotation toward the pitching direction, but there was no difference between right and left hip rotation angles in the disorder group. Hip rotation angle requires hip joint abduction and rotation angles, and therefore the absence of a difference between the right and left sides may indicate a correlation to the disorder.

TRA calls for the recreation of the foot contact phase because during this phase, translational motion shifts to rotational motion. This is an important phase during which potential energy is converted to propulsive force and the lower limb muscle groups of the axis foot that is fixed in place produce maximum momentum [26]. Regarding kinematics, it is performed by eccentric contraction on hip joint external rotation group of muscles and the adductors of the stance leg. However, dysfunctions, such as a restricted range of motion or reduced muscle strength, cause the throwing form to become disordered [27] and as a result, excessive shoulder abduction, reduced step width, and poor shoulder rotation timing occur, causing poor performance [3, 4, 28]. In addition, poor hip and trunk rotation timing and facing the body toward the front during this phase lead to greater stress on the shoulder and elbow joints [5-7], which may lead to throwing disorder. TRA is thought to involve abduction and external rotation of the hip joint on the stance leg side and the angles of flexion, rotation, and abduction of the hip joint on the lead leg side. Moreover, the muscle strength of the adductor on the stance leg side as well as the hamstring and gluteus maximus on the lead leg side may be involved. Our investigation of trunk rotation indicates that, in addition to the aforementioned lower limb range of motion and muscle strength, the trunk rotation range of motion is involved in TRA.

The results of this study indicated significantly lower values for both hip and trunk rotation in the disorder group as compared to the healthy group. These lower limb and trunk dysfunctions are factors that can cause dropped elbow, excessive external rotation of the shoulder joint, and horizontal external rotation during the subsequent late cocking phase, which we believe may lead to a throwing disorder. Subjects with elbow joint disorder in particular showed lower values than healthy subjects for both the hip and trunk rotation angles, which may indicate that the test is a better suited to screen for elbow joint disorder. Further study of this issue with larger numbers of subjects is required.

This assessment method is reproducible, can be easily performed, and assesses complex elements in a short time. Its sensitivity in detecting hip rotation angle disorders is relatively high at 74.8%, which makes it useful as a screening method during medical checkups. It may also be useful as a self-check and partner-check at sporting events. However, because the present study was a pilot study, future detailed assessments of the range of motion for a variety of joints and muscle strength levels using TRA from the point of view of kinetics and dynamics, as well as elucidation of the association between actual throwing motion and TRA are required. Moreover, the investigation of whether improving the range of motion as assessed by TRA can prevent the future onset of shoulder internal impingement and posterior elbow disorders is needed in addition to the development of training methods that improve TRA.

## Conclusion

We devised an assessment method that evaluates complex rotation motions similar to the throwing motion and used it to compare high school pitchers with a throwing disorder to those without (healthy). Our comparison of the healthy and disorder groups indicated that the disorder group had significantly lower values for both the hip and trunk. Disorder was determined by the hip cutoff value of 42.5°, sensitivity of 89.3%, and specificity of 45.6%. The trunk cutoff value was 81°, sensitivity was 63.8%, and specificity was 65%. Our method is reproducible, can be easily used to assess complex elements in a short time, and is useful as a screening method during medical checkups.

## Data Availability

The datasets generated and/or analysed during the current study are not publicly available due to limitations of ethical approval involving the patient data and anonymity but are available from the corresponding author on reasonable request.

## Acknowledgments

We would like to thank Editage (www.editage.com) for English language editing.

